# Reliable detection of eczema areas for fully automated assessment of eczema severity from digital camera images

**DOI:** 10.1101/2022.11.05.22281951

**Authors:** Rahman Attar, Guillem Hurault, Zihao Wang, Ricardo Mokhtari, Kevin Pan, Bayanne Olabi, Eleanor Earp, Lloyd Steele, Hywel C. Williams, Reiko J. Tanaka

## Abstract

Assessing the severity of eczema in clinical research requires face-to-face skin examination by trained staff. Such approaches are resource-intensive for participants and staff, challenging during pandemics, and prone to inter- and intra-observer variation. Computer vision algorithms have been proposed to automate the assessment of eczema severity using digital camera images. However, they often require human intervention to detect eczema lesions and cannot automatically assess eczema severity from real-world images in an end-to-end pipeline.

We developed a new model to detect eczema lesions from images using data augmentation and pixel-level segmentation of eczema lesions on 1345 images provided by dermatologists. We evaluated the quality of the obtained segmentation compared to that of the clinicians, the robustness to varying imaging conditions encountered in real-life images, such as lighting, focus, and blur and the performance of downstream severity prediction when using the detected eczema lesions. The quality and robustness of eczema lesion detection increased by approximately 25% and 40%, respectively, compared to our previous eczema detection model. The performance of the downstream severity prediction remained unchanged.

## INTRODUCTION

Atopic dermatitis (AD, synonym with eczema and atopic eczema) is the most common chronic inflammatory skin disease affecting 15-30% of children and 2-10% of adults worldwide (Langan et al., 2020). Assessment of AD severity can include symptoms such as itching, disease signs such as excoriation, or other aspects of the disease such as control and quality of life. Simple global methods or short symptom and quality of life questionnaires can be used in clinical practice (Leshem et al., 2020).

However, objective assessments of AD severity are increasingly used to assess eligibility for systemic medicines that might be needed for severe disease (National Institute for Health and Excellence, 2018). In research studies such as clinical trials, objective assessment of AD severity is usually considered essential to standardise comparisons and reduce detection biases for interventions that cannot be blinded. The international Harmonising Outcome Measures for Eczema initiative has recently established a core outcome sets for clinical trials for AD that includes objective measurement of signs using EASI (Williams et al., 2022). Objective assessment of AD severity usually requires assessment by trained clinical staff for signs such as redness or lichenification graded from none (=0) to severe (=3) for each sign.

Complete skin examination in a face-to-face environment is desirable for objective assessment of AD severity. However, it is resource-intensive for both the patient/study participant and the trained assessor. Studies that require repeated visits to the clinic for assessments are especially challenging and can contribute to large quantities of missing data. The recent COVID-19 pandemic has placed additional strain on such face-to-face visits. It is also known that inter- and intra-observer variation can be a significant challenge when assessing AD severity objectively (Schmitt et al., 2013).

A form of automated remote assessment of AD severity using digital images is desirable, as it could enable and standardise the remote assessment of AD severity and reduce the inter- and intra-observer variability. However, the methods published to date are often not fully automated to detect AD lesions and assess AD severity. They require manual intervention in their pipeline. For example, Bang et al. (2021) trained and tested a severity assessment algorithm using fixed-size image crops manually prepared to cover only AD areas. Automatic detection of AD lesions in real-world images is required to avoid human intervention and design end-to-end pipelines to assess AD severity from digital camera images.

Pan et al. recently developed a convolutional neural network (CNN)-based computer vision pipeline called EczemaNet (Pan et al., 2020) that first detects AD regions and then assesses the severity of seven disease signs (dryness, erythema, excoriation, cracking, exudation, lichenification, and oedema). The pipeline was trained using real-world images taken with digital cameras in a published clinical trial so that EczemaNet can be used in real-world situations where the images are of different sizes, taken under various imaging conditions (e.g., resolution, lighting, focus, and blur), or include non-AD skin and non-skin background. However, training data for the AD region detection model in EczemaNet was obtained from non-experts, and the model’s detection quality was considered a bottleneck for better severity assessment.

This study aims to develop accurate and robust computer vision algorithms for AD region detection to enable reliable assessment of AD severity from digital images. We use pixel-level AD segmentation data obtained from clinicians that provide granular information on the extent of AD regions in the images. Previously, Nisar et al. (2021) attempted automatic segmentation of AD lesions in 84 images that contain only AD regions and neighbouring pixels, and Son et al. (2021) explored the segmentation and classification of erythema lesions. In comparison, we introduce EczemaNet2 in this study that segments the areas of AD (not only erythema) lesions using 1345 real-world images that include non-AD skin and non-skin background. We will also apply data augmentation techniques and quantify the robustness of AD region detection to varying imaging conditions.

## RESULTS

### Overview of EczemaNet1

The published EczemaNet model (Pan et al., 2020) has two main components (Figure 1): RoI detection model and the severity prediction model. The latter makes probabilistic predictions of seven disease signs of AD for each crop produced by the RoI detection model. The predicted severity of each disease sign is aggregated to produce regional severity scores for the whole image, such as regional versions of Six Area, Six Sign Atopic Dermatitis (SASSAD) score (Berth-Jones, 1996), Three Item Severity Score (TISS) (Wolkerstorfer et al., 1999), and Eczema Area and Severity Index (EASI) (Hanifin et al., 2001).

**Figure 1:**
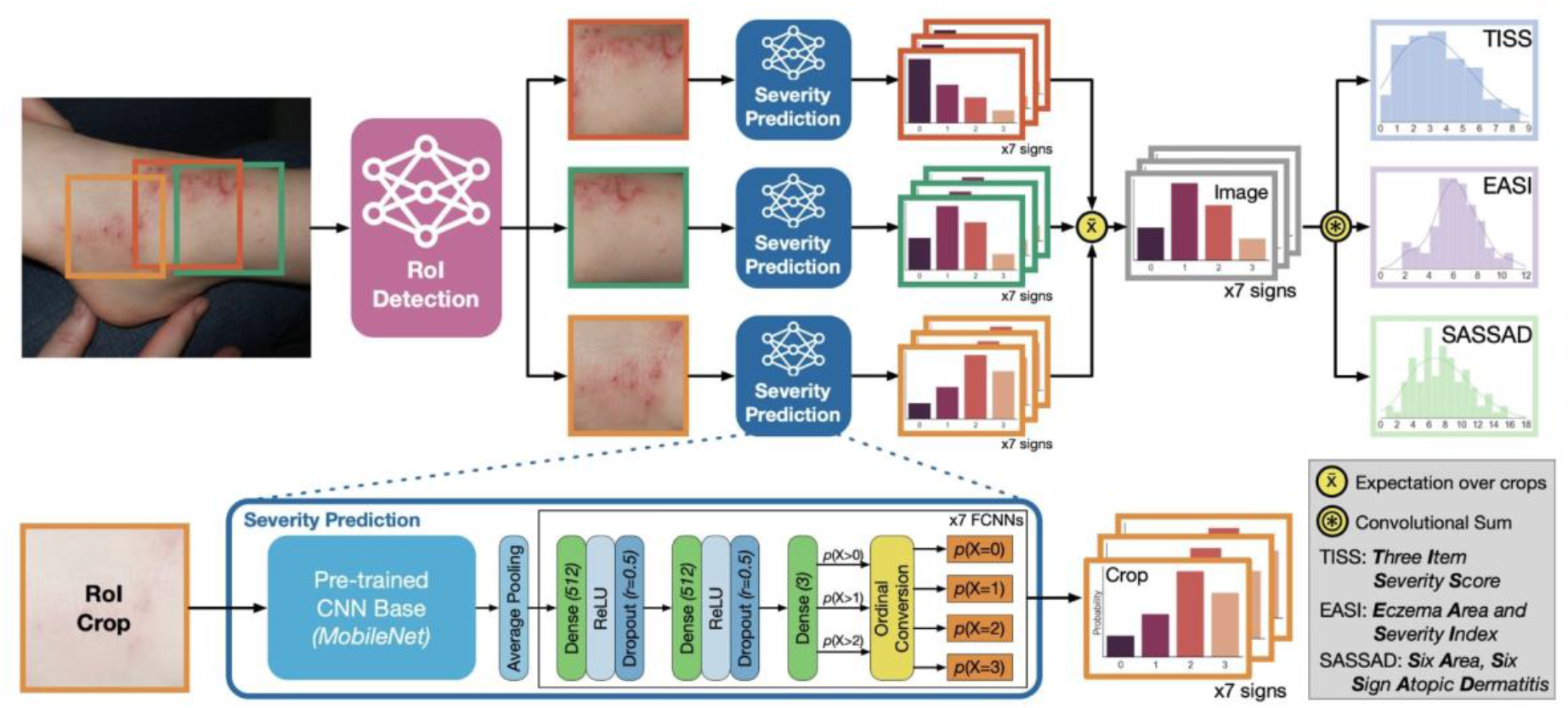
Overview of the EczemaNet1 pipeline, taken from (Pan et al., 2020). The RoI detection model generates AD crops of the input image that contain AD regions. The severity prediction model makes probabilistic predictions of seven disease signs in each crop. The AD severity scores for each disease sign are integrated to give the regional severity scores for the whole image.

The RoI detection model for EczemaNet1 was trained on crops obtained from non-experts. Its outputs are rectangular bounding boxes of the detected AD regions in an image that may also include background pixels and do not provide information about the particular shape of the AD regions, limiting the performance of AD severity assessment in EczemaNet1.

### Pixel-level segmentation data

We used 1345 photos of representative AD regions from 287 AD children aged six months to 16 years collected as a part of the Softened Water Eczema Trial (SWET) (Thomas et al., 2011). In SWET, a clinical staff took photos and recorded the corresponding severity of seven disease signs (dryness, erythema, excoriation, cracking, exudation, lichenification, and oedema) from none (=0) to severe (=3) assessed in person. The photos vary in resolution and image quality, including focus, lighting, and blur.

Four dermatologists delineated (“segmented”) AD regions at the pixel-level in the 1345 images (307 images by BO, 308 images by EE, 308 images by LS and 422 images by HCW). This pixel-level AD segmentation data offers a finer resolution of AD regions than the 1748 AD crops used to train the RoI detection model in EczemaNet1, where the AD crops were manually extracted by three non-medical students rather than by dermatologists. In addition to the pixel-level AD segmentation, a non-medical student (RM) provided skin segmentation at the pixel level, resulting in pixel-level segmentation masks of background, non-AD skin and AD skin (Figure 2).

**Figure 2:**
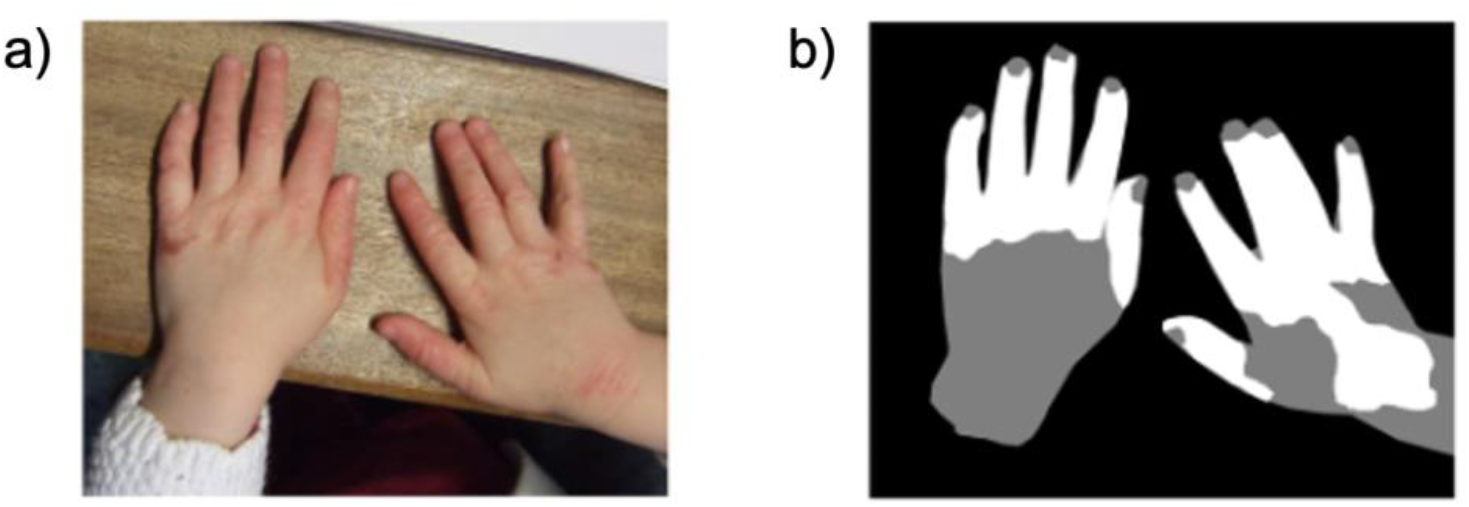
Illustration of pixel-level segmentation masks. (a) A sample image and (b) the corresponding pixel-level segmentation masks of background (black), non-AD skin (grey) and AD skin (white).

### A new RoI detection model using pixel-level segmentation for EczemaNet2

We present EczemaNet2, for which we modified the RoI detection model of EczemaNet1 with a standard pixel-level segmentation U-Net (Ronneberger et al., 2015). We trained two U-Nets with pixel-level segmentation masks: one produces skin segmentation masks, and another produces AD segmentation masks, respectively, for every image (Figure 3(a)). We also added three sequential post-processing steps to extract square crops (Figure 3(b)) required for the subsequent severity prediction model. The first step was the border following to generate rectangular crops by topological structural analysis of binary images with the border representation of AD regions obtained by a morphological operator (Suzuki and others, 1985). The second step was the square cropping to resize and extract square crops of the same dimension based on the aspect ratio of the rectangular crops. It avoids image distortion and changes in the appearance and proportion of AD regions that occurred in the resizing step of EczemaNet1, in which image crops were squashed or stretched. The final post-processing step was to add surrounding non-AD skin pixels to the AD skin pixels in the square crops. This step was applied only when both skin and AD segmentation masks were available.

**Figure 3:**
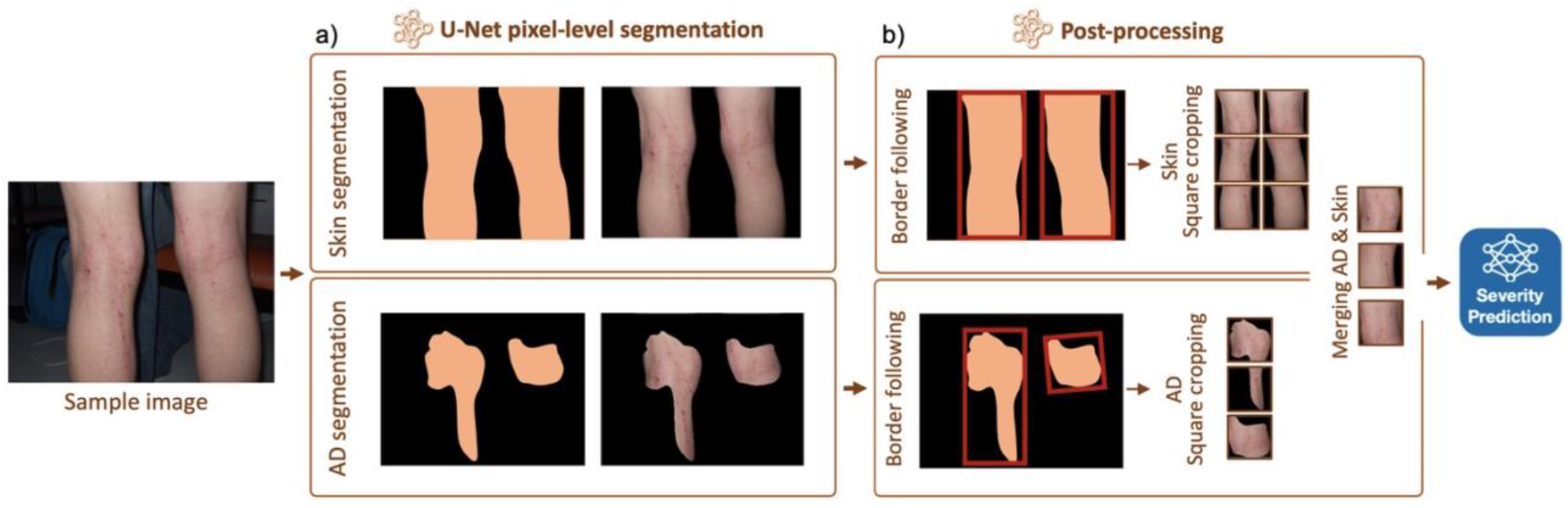
Overview of the RoI detection part of EczemaNet2. (a) U-Net pixel-level segmentation for skin or AD and (b) post-processing steps to produce crops that are inputs for the subsequent severity prediction model. Merging non-AD skin pixels with AD pixels is applied when both AD and Skin segmentation are available.

### Systematic evaluation of the quality and robustness of RoI detection

We compared the performance of the region-of-interest (RoI) models with various configurations of training data and data augmentation (Table 1). The performance was evaluated in terms of the quality and the robustness of RoI detection, two essential aspects of computer vision methods to be used in clinical practices. The same data was used for a consistent and fair comparison of all models, with a split ratio of 6:2:2 for training, validation, and test sets. We also investigated whether the accuracy of the downstream severity prediction task changed.

**Table 1:** Summary of the AD detection pipelines.

| Configuration name | Pipeline | RoI model | Training data | Data augmentation |
| --- | --- | --- | --- | --- |
| Base | EczemaNet1 | A faster R-CNN | AD crops | No |
| Skin | EczemaNet2 | A U-Net | Skin segmentation masks | No |
| AD |  |  | AD segmentation masks | No |
| AD/Skin |  | 2 U-Nets | AD & skin segmentation masks | No |
| AD/Skin+Pix2Pix |  |  | AD & skin segmentation masks | Pix2Pix |
| AD/Skin+DA |  |  | AD & skin segmentation masks | Traditional |

Firstly, we evaluated the quality of RoI detection using the F1 score and precision for each image based on the classification of each pixel into AD region or not in all relevant crops (Figure 4(a)). Pixel-level segmentation of AD regions obtained from four dermatologists was used as a reference.

**Figure 4:**
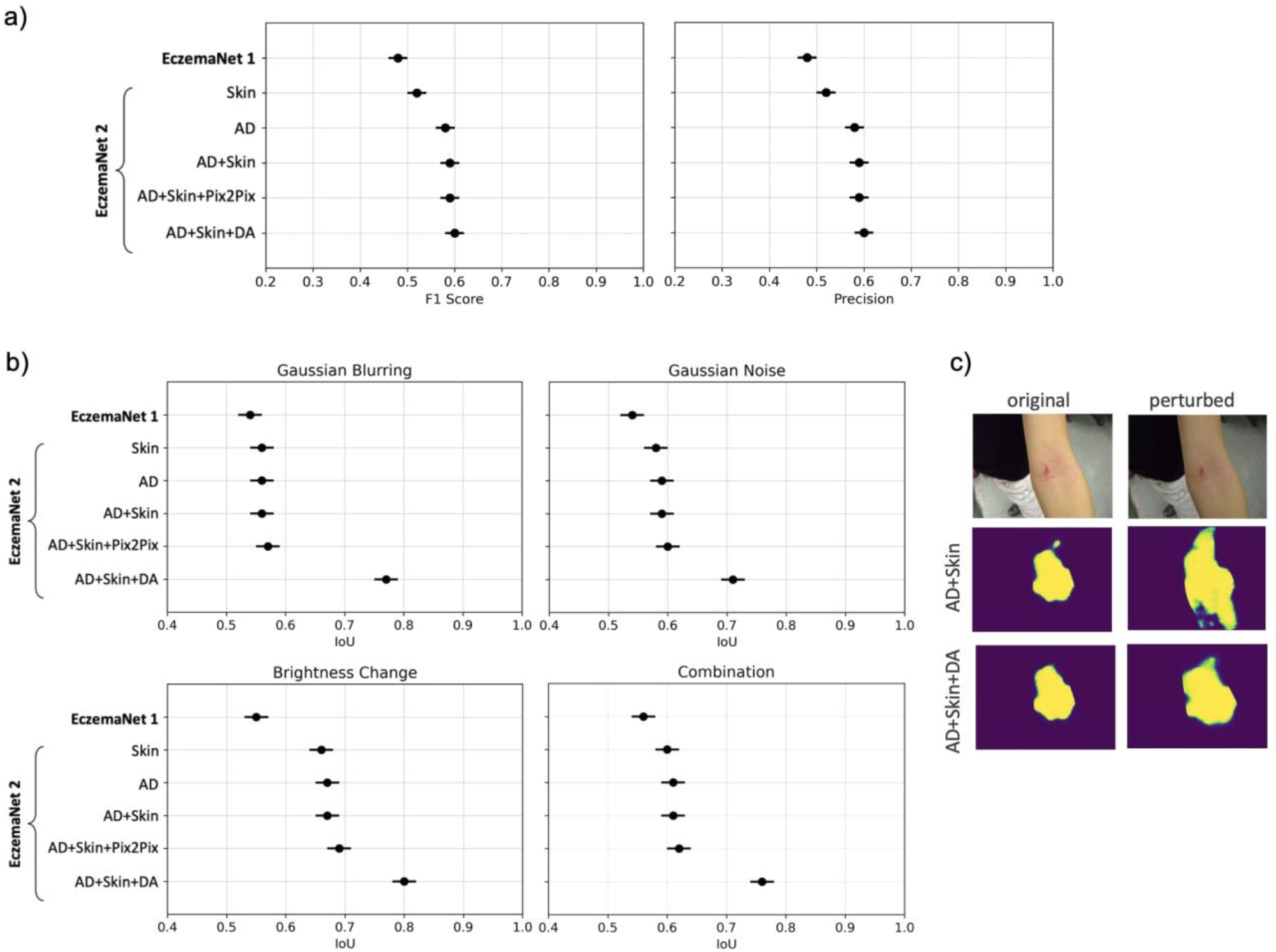
Quality and robustness of RoI detection. (a) Quality measured by F1 score and precision (mean ± SE, the higher, the better). (b) Robustness of RoI detection against image perturbations measured by IoU between predictions made from unperturbed vs perturbed images (the higher the better), for different perturbations (Gaussian blurring, Gaussian noise, brightness change, and combinations). (c) Example AD segmentation of unperturbed and perturbed images by the RoI models trained with an augmented dataset (“AD+Skin+DA”) and with the non-augmented dataset (“AD+Skin”). “AD+Skin+DA” generates consistent masks even in the presence of image perturbations, but not “AD+Skin”.

The RoI detection model of EczemaNet2 trained on pixel-level skin segmentation masks (“Skin” in Table 1 and Figure 4(a)) achieved a better detection quality than the RoI detection model of EczemaNet1. The RoI detection model of EczemaNet2 trained on pixel-level AD segmentation masks (“AD”) achieved a slightly better quality than that on skin segmentation masks. This was expected as training the model with skin segmentation masks leads to many false positives because all predicted skin pixels are considered AD. The RoI quality was not improved by adding surrounding non-AD skin pixels to AD skin segmentation masks (“AD+Skin”) and data augmentation with traditional methods (“AD+Skin+DA”) and Pix2Pix (“AD+Skin+Pix2Pix”). Despite the improvement in AD detection brought by EczemaNet2 from EczemaNet1, the detection performance remains moderate, with an average precision and F1 score of approximately 60% across images.

Next, we evaluated the robustness of RoI detection, i.e., the sensitivity of the model’s predictions to perturbations in the model’s inputs, by quantifying the similarity between the AD regions detected from unperturbed vs perturbed input images using the intersection over union (IoU). The perturbations applied are similar to those applied for data augmentation (Table 2), including blurring (e.g. due to incorrect focus), brightness changes (e.g. due to incorrect exposure), noise (e.g. due to poor lighting conditions) and their combinations that could realistically occur in images taken with a smartphone camera.

**Table 2:** Traditional data augmentation methods applied to create an augmented dataset.

| Method | Range: Increment size | # Generated images |
| --- | --- | --- |
| Blur | [2.5, 12.5] : 2.5 | 5X |
| Noise | [0.01, 0.05] : 0.01 | 5X |
| Brightness | [0.5, 1.5] : 0.2 | 6X |
| Zoom | [0.25, 1.75] : 0.25 | 6X |
| Rotation | {-90, 90} | 2X |
| Flip | {horizontal, vertical} | 2X |
| Mix | Combination of two randomly chosen methods | 2X |

RoI detection in EczemaNet2 is more robust than EczemaNet1 in terms of IoU for all perturbations considered (Figure 4(b)). Data augmentation of training data using traditional methods (“DA”) improved the robustness of RoI detection (Figure 4(c)), but data augmentation by Pix2Pix did not. These results suggest that complex augmentation methods are not necessarily required to improve the robustness of AD detection models. The IoU of the most robust configuration (“AD+Skin+DA”) still did not reach a perfect score of 1, suggesting that the AD segmentation remains sensitive to irrelevant features of the images.

### Accuracy of severity prediction

Finally, we investigated the impact of the different RoI model configurations on the downstream severity prediction task (Figure 5). We conducted 10-fold cross-validation with a 90:10 train/test split, stratified on patients, and computed the root mean square error (RMSE) of the mean prediction across test images.

**Figure 5:**
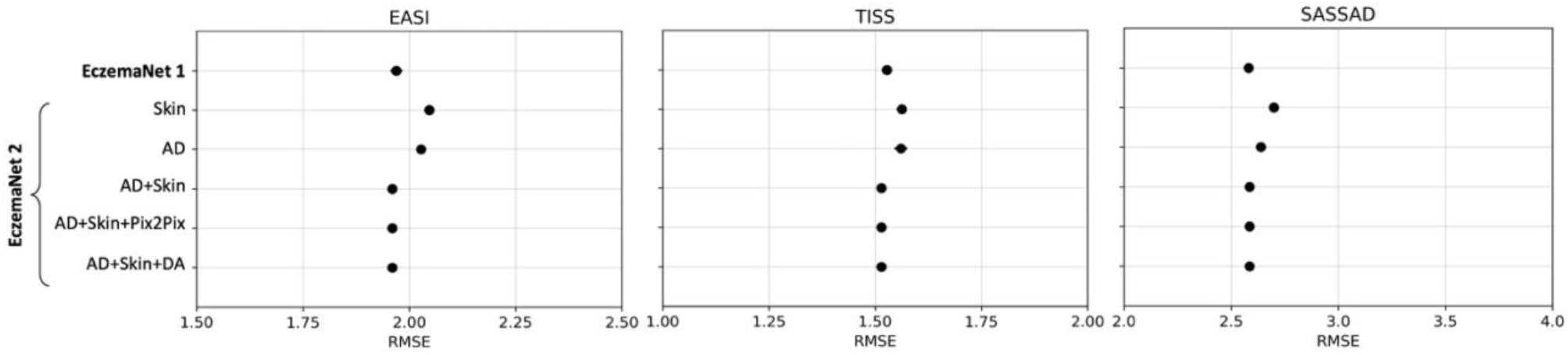
Accuracy of severity prediction evaluated with RMSE. Points show the mean ± SE over 10-fold cross-validation for the regional EASI (in [0, 12]), TISS (in [0, 9]), and SASSAD (in [0, 18]).

We evaluated the performance in predicting the regional scores for EASI (erythema, excoriation, lichenification, and oedema), TISS (erythema, excoriation, and oedema), and SASSAD (cracking, dryness, erythema, excoriation, exudation, and lichenification). The RoI detection models in EczemaNet2 trained with AD or skin segmentation masks (“Skin”, “AD”) achieved a better performance than that in EczemaNet1 (Figure 4). It resulted in a slightly higher RMSE for AD severity assessment (Figure 5), although the practical significance of the difference in RMSE is debatable. Merging AD segmentation masks with the neighbouring skin pixels (“AD+Skin”) provided the downstream severity model with informative and discriminative pattern features between the healthy skin and the lesion (e.g. brightness colour, gradient and texture changes) and led to marginal improvement in the accuracy of severity prediction compared to EczemaNet1 and EczemaNet2 (“AD” or “Skin”). Data augmentation, either with traditional methods (“DA”) or Pix2Pix, did not impact the average predictive performance.

## DISCUSSION

### Main findings

This study presents EczemaNet2, in which we propose a new algorithm to detect AD regions (region-of-interest, RoI, Figure 3) to enable reliable assessment of AD severity from digital camera images. It builds on and improves previously pusblished EczemaNet1 (Figure 1), capable of detecting AD regions from camera images and subsequently assessing the severity of seven AD disease signs without any manual intervention. The RoI model of EczemaNet2 was trained on pixel-level AD segmentation masks provided by four dermatologists (Figure 2). EczemaNet2 could detect and extract AD regions from digital images more accurately than EczemaNet1 (Figure 4(a)). Data augmentation for the training set boosted the model’s robustness to poor imaging conditions and external noise that is often found in real-life images (Figure 4(b)(c)).

### Strengths and limitations of this study

The originality of this study lies in thorough and systematic evaluation of the performance and robustness of AD detection algorithms, using AD, AD+Skin, and Skin, together with data augmentation (Pix2Pix and traditional data augmentation methods). Study strengths include thorough experimentations to evaluate the contribution of different model improvements in terms of detection quality, robustness to poor imaging conditions and severity prediction performance. By improving the detection quality of the RoI model in EczemaNet2, we improve the interpretability of the entire pipeline, as we can be more confident that the severity of AD is assessed from relevant image features. Interpretability of models is relevant to patients’ “right to an explanation” for automated decision-making, highlighted in existing regulations such as the European General Data Protection Regulation (Goodman and Flaxman, 2017). We used privileged information with 1345 pixel-level segmentation masks provided by dermatologists and further expanded the dataset with data augmentation techniques. Data augmentation improved the robustness of the RoI model, which is essential to maintain the users’ trust in automated AD severity assessment and ensure the models provide consistent predictions without being sensitive to perturbations in input images.

We recognise that AD segmentation masks may be unreliable, as a poor agreement among dermatologists was found in detecting AD regions in digital images (Hurault et al., 2022). Poor inter-rater reliability of AD segmentation could explain why the detection accuracy of AD regions remains low in EczemaNet2 (Figure 5). It could further explain why the U-Net did not perform much better for AD detection when it was trained to delineate AD regions as opposed to delineating skin and why merging AD segmentation with surrounding skin pixels achieved only slightly better performance in severity prediction. We believe skin segmentation may be a reasonable alternative to AD segmentation that would require specialist labelling whose quality may be debatable. By definition, skin segmentation does not identify AD lesions. Still, it may sufficiently restrict the inputs to the severity assessment model without excluding potentially informative regions in the images (therefore achieving perfect sensitivity/recall), assuming the images contain a priori representative sites of AD. It could nonetheless be interesting to explore AD segmentation algorithms that can deal with noisy segmentation labels (Karimi et al., 2020).

### Implications for clinical practice and research

This study proposed a robust method to detect AD regions from digital images while highlighting the challenges of this endeavour. Accurate and robust detection of AD regions is necessary for developing end-to-end pipelines that automatically assess AD severity from real-world digital images. While there is considerable promise for remote assessment of AD, the performance of the downstream severity prediction remained unchanged in EczemaNet2, highlighting the difficulty of assessing AD severity from real-world digital images. We believe that collecting more and better-quality data (images and labels) would surpass the gains in performance from using cleverer algorithms. In particular, we emphasise the importance of ensuring that the training dataset covers images for various skin tones to limit skin colour bias (Daneshjou et al., 2021).

## MATERIAL AND METHODS

### Data augmentation

Data augmentation refers to the artificial expansion of a dataset by adding synthetic data to increase its size and variation (Shorten and Khoshgoftaar, 2019). Data augmentation is often applied to mitigate the adverse effects of training on a small dataset, such as overfitting to training data, producing an improved and more robust model that can better generalise to unseen data.

We created an augmented dataset to train the segmentation model of EczemaNet2. We applied Gaussian blurring and Gaussian noise to the images, brightened, darkened, zoomed in and out, rotated and flipped the images. The range and increment size of the scaling parameters were chosen to ensure the augmented dataset includes various images that may be encountered during image acquisition realistically (Table 2). Those values determined the number of generated images for each augmentation method. For example, five new images were produced by blurring every image in the original dataset with five levels of blurriness. We also applied a combination of two randomly chosen augmentation methods to synthesise two images per original image (Mix in Table 2). The final augmented dataset contains all the synthesised images (28 images in total per original image) and the original images.

We also created another augmented dataset by applying a more complex augmentation method to compare the effects of the data augmentation method on the performance of RoI detection. We used Pix2Pix (Isola et al., 2017), which is based on conditional Generative Adversarial Networks (Mirza and Osindero, 2014) and can generate synthetic images from arbitrary segmentation masks (i.e., images with non-AD/AD/background labels in our case) that describe the precise location of RoI in images.

### Model implementation and training

All algorithms tested in this study were developed using libraries and scripts in Python 3.7 and TensorFlow 1.15. The RoI model of EczemaNet2 was trained using Adam for optimising the cross-entropy loss function through 50 epochs with a learning rate of 0.0001 and batch size of 2 samples, all of which were determined empirically.

## Data Availability

Source code is publicly available at https://github.com/Tanaka-Group/EczemaNet2.

## REFERENCES

Bang, C.H., Yoon, J.W., Ryu, J.Y., Chun, J.H., Han, J.H., Lee, Y.B., Lee, J.Y., Park, Y.M., Lee, S.J., Lee, J.H., 2021. Automated severity scoring of atopic dermatitis patients by a deep neural network. Sci Rep 11, 1–8.

Berth-Jones, J., 1996. Six Area, Six Sign Atopic Dermatitis (SASSAD) seventy score: A simple system for monitoring disease activity in atopic dermatitis. British Journal of Dermatology, Supplement 135, 25–30. https://doi.org/10.1111/j.1365-2133.1996.tb00706.x

Daneshjou, R., Smith, M.P., Sun, M.D., Rotemberg, V., Zou, J., 2021. Lack of Transparency and Potential Bias in Artificial Intelligence Data Sets and Algorithms: A Scoping Review. JAMA Dermatol 157, 1362–1369. https://doi.org/10.1001/JAMADERMATOL.2021.3129

Goodman, B., Flaxman, S., 2017. European Union Regulations on Algorithmic Decision Making and a “Right to Explanation.” AI Mag 38, 50–57.

Hanifin, J.M., Thurston, M., Omoto, M., Cherill, R., Tofte, S.J., Graeber, M., Evaluator Group, T.E., 2001. The eczema area and severity index (EASI): assessment of reliability in atopic dermatitis. Exp Dermatol 10, 11–18. https://doi.org/10.1034/j.1600-0625.2001.100102.x

Hurault, G., Pan, K., Mokhtari, R., Olabi, B., Earp, E., Steele, L., Williams, H.C., Tanaka, R.J., 2022. Detecting eczema areas in digital images: an impossible task? medRxiv. https://doi.org/10.1101/2022.03.03.22271780

Isola, P., Zhu, J.-Y., Zhou, T., Efros, A.A., 2017. Image-to-image translation with conditional adversarial networks, in: Proceedings of the IEEE Conference on Computer Vision and Pattern Recognition. pp. 1125–1134.

Karimi, D., Dou, H., Warfield, S.K., Gholipour, A., 2020. Deep learning with noisy labels: Exploring techniques and remedies in medical image analysis. Med Image Anal 65, 101759.

Langan, S.M., Irvine, A.D., Weidinger, S., 2020. Atopic dermatitis. The Lancet 396, 345– 360. https://doi.org/10.1016/s0140-6736(20)31286-1

Leshem, Y.A., Chalmers, J.R., Apfelbacher, C., Furue, M., Gerbens, L.A.A., Prinsen, C.A.C., Schmitt, J., Spuls, P.I., Thomas, K.S., Williams, H.C., Simpson, E.L., 2020. Measuring atopic eczema symptoms in clinical practice: The first consensus statement from the Harmonising Outcome Measures for Eczema in clinical practice initiative. J Am Acad Dermatol 82, 1181–1186. https://doi.org/10.1016/J.JAAD.2019.12.055

Mirza, M., Osindero, S., 2014. Conditional Generative Adversarial Nets. arXiv preprint arXiv:1411.1784.

National Institute for Health, Excellence, C., 2018. Dupilumab for treating moderate to severe atopic dermatitis.

Nisar, H., Tan, Y.R., Ho, Y.K., 2021. Segmentation of Eczema Skin Lesions Using U-Net, in: 2020 IEEE-EMBS Conference on Biomedical Engineering and Sciences (IECBES). pp. 362–366.

Pan, K., Hurault, G., Arulkumaran, K., Williams, H., Tanaka, R.J., 2020. EczemaNet: Automating Detection and Assessment of Atopic Dermatitis, in: Machine Learning In Medical Imaging. pp. 220–230.

Ronneberger, O., Fischer, P., Brox, T., 2015. U-net: Convolutional networks for biomedical image segmentation, in: International Conference on Medical Image Computing and Computer-Assisted Intervention. pp. 234–241.

Schmitt, J., Langan, S., Deckert, S., Svensson, A., von Kobyletzki, L., Thomas, K., Spuls, P., 2013. Assessment of clinical signs of atopic dermatitis: A systematic review and recommendation. Journal of Allergy and Clinical Immunology 132, 1337–1347. https://doi.org/10.1016/j.jaci.2013.07.008

Shorten, C., Khoshgoftaar, T.M., 2019. A survey on image data augmentation for deep learning. J Big Data 6, 1–48.

Son, H.M., Jeon, W., Kim, J., Heo, C.Y., Yoon, H.J., Park, J.-U., Chung, T.-M., 2021. AI-based localization and classification of skin disease with erythema. Sci Rep 11, 1–14.

Suzuki, S., others, 1985. Topological structural analysis of digitized binary images by border following. Comput Vis Graph Image Process 30, 32–46.

Thomas, K.S., Koller, K., Dean, T., o’Leary, C.J., Sach, T.H., Frost, A., Pallett, I., Crook, A.M., Meredith, S., Nunn, A.J., others, 2011. A multicentre randomised controlled trial and economic evaluation of ion-exchange water softeners for the treatment of eczema in children: the Softened Water Eczema Trial (SWET). Health Technol Assess (Rockv) 15, 1–156.

Williams, H.C., Schmitt, J., Thomas, K.S., Spuls, P.I., Simpson, E.L., Apfelbacher, C.J., Chalmers, J.R., Furue, M., Katoh, N., Gerbens, L.A.A., others, 2022. The HOME Core outcome set for clinical trials of atopic dermatitis. Journal of Allergy and Clinical Immunology.

Wolkerstorfer, A., de Waard Van Der Spek, F.B., Glazenburg, E.J., Mulder, P.G.H., Oranje, A.P., 1999. Scoring the Severity of Atopic Dermatitis: Three Item Severity Score as a Rough System for Daily Practice and as a Pre-screening Tool for Studies. Acta Derm Venereol 79, 356–359.

